# Phase I dose escalation of the Exportin 1 inhibitor, Selinexor, in combination with chemoradiation in patients with newly diagnosed glioblastoma

**DOI:** 10.64898/2026.06.25.26356263

**Authors:** Peter Mathen, Huma Chaudhry, Megan Mackey, Theresa Cooley-Zgela, Matthew Masciocchi, Bo Li, Erich Huang, Jing Wu, DeeDee Smart, Andra Krauze, Kevin Camphausen

**Affiliations:** Radiation Oncology Branch, NCI, NIH, Bethesda, MD 20892; Neuro-Oncology Branch, NCI, NIH, Bethesda, MD 20892; Biometric Research Program, NCI, NIH, Bethesda, MD 20892

**Keywords:** GBM, radiation, temozolomide, Selinexor, radiosensitizer

## Abstract

**Purpose:** Glioblastoma (GBM) remains associated with poor outcomes, with most recurrences occurring within the high-dose radiation field suggesting persistent radioresistance. Exportin 1 (XPO1) inhibition with Selinexor has demonstrated radiosensitizing effects in preclinical models. We conducted a phase I trial to evaluate the safety, tolerability, and preliminary efficacy of Selinexor in combination with standard chemoradiation for newly diagnosed GBM.

**Methods:** This investigator-initiated phase I dose-escalation trial (3+3 design) enrolled adults with newly diagnosed GBM or gliosarcoma. Patients received standard radiotherapy (60 Gy in 30 fractions) with concurrent temozolomide and escalating doses of Selinexor. Three dose levels were evaluated: 80 mg weekly (weeks 1, 2, 4, 5); 60 mg twice weekly (weeks 1, 2, 4, 5); and 60 mg twice weekly (weeks 1-6) throughout radiotherapy. The primary endpoint was determination of the maximum tolerated dose (MTD) based on dose-limiting toxicities (DLTs). Secondary endpoints included progression-free survival (PFS), overall survival (OS), patterns of failure, and patient-reported outcomes (MDASI-BT).

**Results:** Eleven patients were enrolled. Median age was 58 years, and median KPS was 90. The MTD was established at Selinexor 60 mg twice weekly during weeks 1, 2, 4, and 5 of chemoradiation. Dose level 3 exceeded the MTD with two DLTs. Treatment compliance was high, with minimal missed radiotherapy fractions. Median PFS was 15.9 months (95% CI, 6.2–28.5), and median OS was 17.4 months (95% CI, 14.1–not reached). Most recurrences were central (5/6 evaluable patients). Notably, multiple cases of delayed pseudoprogression were observed at 5, 9, 10, and 23 months post-radiotherapy. Patient-reported symptom burden remained stable over time.

**Conclusions:** Selinexor can be safely combined with standard chemoradiation in patients with newly diagnosed GBM, with an MTD of 60 mg twice weekly during select treatment weeks. Preliminary efficacy signals and an increased incidence of delayed pseudoprogression suggest a potential radiosensitizing effect. These findings support further investigation of Selinexor in larger, prospective studies.

## Introduction

Despite advances in surgical and radiation techniques, glioblastoma (GBM) remains associated with poor prognosis. Median survival for newly diagnosed patients is approximately 16 months^1^ and most recurrences occur within the high-dose radiation field^2^, highlighting the need for improved local therapies. Extent of resection is associated with survival, and multiple randomized trials have established postoperative radiotherapy as beneficial^3–6^. However, attempts to improve outcomes through radiation dose escalation, altered fractionation, or brachytherapy have not demonstrated meaningful survival gains and are often associated with increased toxicity^7–9^. As established in the Phase III EORTC/NCIC trial the addition of temozolomide (TMZ) to radiotherapy, improves both overall and progression free survival and remains the standard of care ^10^. However, because TMZ was administered both concurrently with and following radiotherapy, its role as a radiosensitizer remains uncertain ^11–13^. Although GBM cells *in situ* demonstrate migratory and invasive properties, the typical recurrence pattern is within the high dose radiation field indicative of radioresistance. Thus, the effectiveness of GBM therapy could be improved with the addition of radiosensitizing agents^14^.

Exportin 1 (XPO1) is a key mediator of nuclear export and is responsible for the transport of multiple RNAs and tumour suppressor proteins ^15,16^. XPO1 is overexpressed in GBM and is associated with poor clinical outcomes, supporting its role as a therapeutic target. Selinexor (KPT-330), an oral selective inhibitor of nuclear export, blocks XPO1 function and restores nuclear retention of tumour suppressor proteins and ribosomal (rRNAs), small nuclear (snRNAs), and a certain subset of messenger RNAs (mRNAs)^17–19^. Preclinical studies have demonstrated antitumour activity of Selinexor in GBM models, including effective blood–brain barrier penetration ^20^. In addition, Selinexor has shown to enhance of radiosensitivity in glioblastoma stem-like and established cell lines *in vitro* and *in vivo* ^21^ an effect correlated with the inhibition of the nuclear export of ribosomal RNA leading to an effect on gene translation.

To date, Selinexor has not been evaluated in combination with standard chemoradiation in patients with newly diagnosed GBM or gliosarcoma. We conducted a phase I dose escalation trial, to determine the maximum tolerated dose (MTD) of Selinexor in combination with standard of care chemoradiotherapy (CRT). Herein, we report that Selinexor can be given safely with the combination of temozolomide and radiotherapy in patients with a diagnosis of GBM.

## Methods

### Patient Selection

This investigator-initiated prospective study was a phase I dose escalation trial to determine the maximum tolerated dose (MTD) of Selinexor in combination with standard dose chemoradiotherapy (CRT) and was registered at www.clinicaltrials.gov (NCT04216329). The protocol and informed consent were approved by the institutional review board of the National Institutes of Health, USA, and written informed consent was obtained from all patients.

Adult patients (age >18) with newly diagnosed glioblastoma or gliosarcoma who were able to receive definitive external beam radiation, temozolomide and Selinexor were eligible for this trial. All patients were required to have a KPS ≥ 70 and be ≤ 8 weeks since surgical resection. Patients who were pregnant or breastfeeding, or who had previously been treated with chemotherapy, radiation therapy, gliadel wafers or tumour treating fields, or had pre-existing known or suspected radiation sensitivity syndromes or those with known coagulation problems, liver dysfunction, significant gastrointestinal issues, or active uncontrolled or suspected infections were excluded.

### Study Design

The trial design (3+3 design) included three dose levels. Each dose level included a minimum of three patients (**Figure 1**). If no patient treated at a given dose level experienced a dose-limiting toxicity (DLT), patients were entered at the next dose level. If one of the three patients experienced toxicity, then a minimum of three additional patients were enrolled at that dose level. If two or more patients in any dose level experienced DLT, the MTD was considered to have been exceeded, and escalation was stopped. The initial dose level was 80 mg/week given on weeks 1, 2, 4 and 5 given with standard of care chemoradiation. This dose was then increased to 60 mg given twice a week (week 1, 2, 4, and 5) for the second dose level, and finally 60 mg given twice a week of treatment (week 1-6). Prophylactic concomitant treatments with a 5-HT3 antagonist and olanzapine 5 mg were provided 3 days prior to and during treatment with each dose of Selinexor. Temozolomide was given at the start of radiation and administered orally with at a daily dose of 75 mg/m^2^ during the radiation treatment. Beginning 1-month post-RT adjuvant temozolomide was given as per standard of care (150-200mg/m^2^) for a minimum of 6 months.

**Figure 1:**
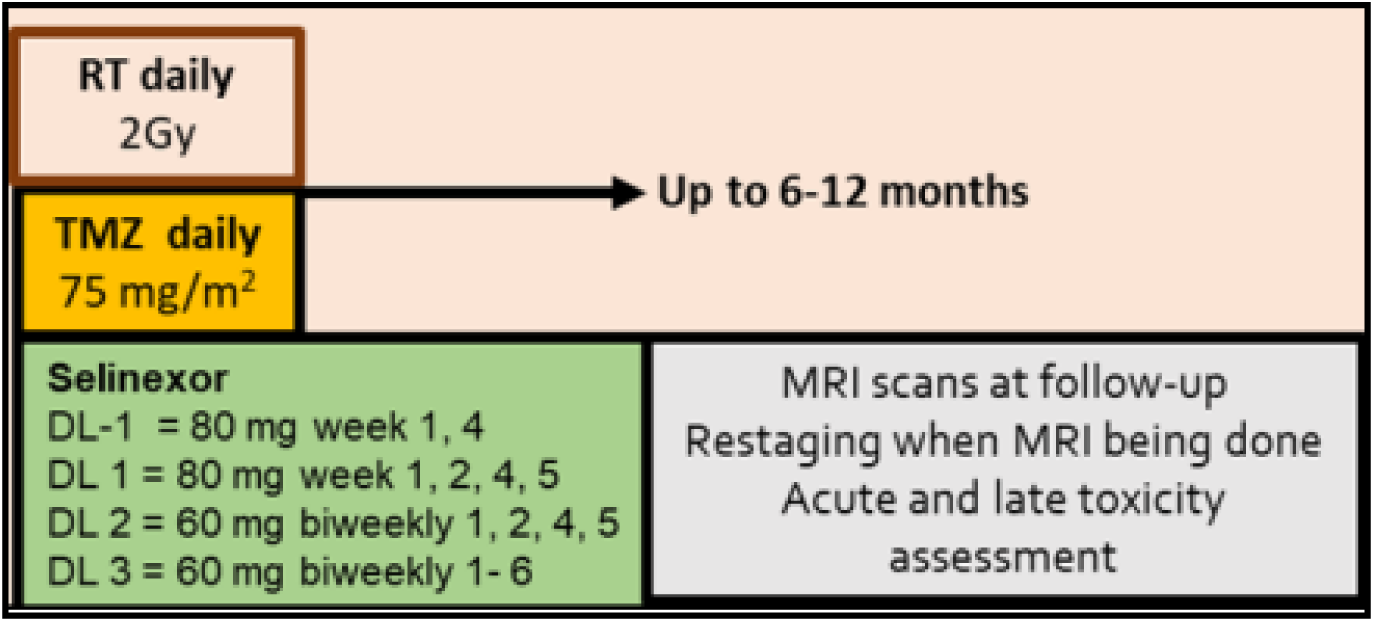
Clinical trial treatment schema. Selinexor was administered concurrently with RT. No adjuvant Selinexor was given. RT = radiation therapy; TMZ = temozolomide; DL = dose level.

Radiation was delivered using intensity-modulated RT (IMRT) and volumetric-modulated arc therapy (VMAT) techniques in 2 Gy fractions to a total dose of 60 Gy. Either EORTC or RTOG treatment volumes were allowed. Gross tumour volume (GTV) for EORTC volumes and GTV2 for RTOG volumes were defined as the enhancing tumour volume seen on T1 Gadolinium MRI sequence images or as postoperative resection cavity, if no residual enhancing tumour was noted. GTV1 for RTOG volumes was the GTV2 volume plus surrounding edema (hyperintensity on T2 or FLAIR MRI scans). A clinical target volume (CTV) expansion of 20 mm and a planning target volume (PTV) expansion of 3-5 mm were used. Details regarding volumes treated and specific dose limits are described in **Supplemental Table 1**.

### Assessments

Clinical response was evaluated using neurological examination and contrast-enhanced MRI. Brain MRI was performed at baseline, one month after completing CRT, and every two months thereafter for two years. Patients with stable disease at that time were imaged with MRI scans every three months for the third year. Imaging response was evaluated by standard Response Assessment Neuro-Oncology (RANO) criteria^22^. Neurologic decline without radiographic evidence of tumour was designated as treatment-related toxicity. Survival duration is determined by the interval from initiation of treatment on the protocol to the date of death. Pattern of failure (POF) was documented as per protocol endpoints (POF_p_). Adverse events (AEs) were graded according to National Cancer Institute Common Terminology Criteria for Adverse Events (NCI-CTCAE) version 5.0. Late radiation related side effects were graded on the RTOG scale.

### Patient-reported Outcomes

Patients self-reported symptom burden using patient-reported questionnaires. Symptom burden was assessed using the MD Anderson Symptom Inventory for Brain Tumors (MDASI-BT). These questionnaires were filled out by patients at baseline, completion of treatment (COT), and follow-up intervals.

### Statistical considerations

The primary objective of this trial was to determine the safety, toxicity profile, DLT, and maximum tolerated Selinexor dose (MTD) in patients with glioblastoma in combination with CRT. The DLT timeframe was defined in the protocol from entry onto protocol to 30 days post-RT. The secondary objectives included the estimation of median progression-free survival (PFS) and overall survival (OS), assessment of late toxicity secondary to Selinexor/CRT, and determination of the impact of Selinexor/CRT on the patient’s HRQOL symptom burden.

## Results

### Patient Characteristics

Eleven patients with primary grade IV glioma, were enrolled between July 2020 and July 2023, and underwent treatment per the schema shown in **Figure 1**. All patients received concurrent daily temozolomide and radiation for six weeks. Patients’ baseline characteristics and treatment parameters are listed in **Table 1**. Seven patients were female, the median age was 58 (range 48-71), and the median KPS was 90 (range 80-100). Ten patients had grade IV glioma, with the sole remaining patient having grade IV gliosarcoma. Extent of surgery was GTR (n=3), STR (n=7) and biopsy (n=1). Four patients had tumours in the temporal lobe, three in the frontal lobe, two in the parietal lobe and the remaining 2 were multi-lobar. The methylation status of the tumours was 45% methylated and 100% IDH wild-type. The median GTV at treatment was 24.2 cc (range 7.5-40.7 cc), the CTV was 162.7 cc (range 75.7-300.2 cc), and the PTV was 222.3 cc (range 116.1-388.2 cc) **Supplemental Table 1**.

**Table 1:** Selinexor study pretreatment characteristics. KPS = Karnofsky performance score, GTR = gross total resection, STR = sub-total resection, Bx = biopsy, MGMT = methyl-guanine methyl transferase, IDH = isocitrate dehydrogenase, TMZ = temozolomide.

| Gender | Number/Percent (%) |
| --- | --- |
| Male | 4(36) |
| Female | 7(64) |
| Age (y) |  |
| median (range) | 58 (48-71) |
| KPS |  |
| median (range) | 90 (80-100) |
| Resection |  |
| GTR | 3 |
| STR | 7 |
| Bx | 1 |
| Tumor Location |  |
| Frontal | 3 |
| Parietal | 2 |
| Temporal | 4 |
| MGMT |  |
| Positive | 5 |
| Negative | 6 |
| IDH |  |
| WT | 11 |
| Radiation |  |
| Completed | 10 |
| Concurrent TMZ |  |
| Completed | 6 |
| Adjuvant TMZ (6m) |  |
| Completed | 8 |

### Toxicity and MTD

At the time of trial initiation, the standard FDA approved dosing schedule for Selinexor was once per week, therefore Selinexor was dosed only once per week in the first dose level. During the first dose level, it was determined that twice weekly dosing was also safe, thus, our protocol was amended and twice weekly dosing for dose levels 2 and 3. At dose level 1, all patients completed therapy without Selinexor related grade 3 or 4 toxicity as reported in **Table 2**. At dose level 2, one patient had five grade 3 or 4 toxicities. Specifically, two episodes of grade 3 *neutrophil count decreased*, one grade 3 *platelet count decreased* and two episodes of grade 3 *white blood cell count decreased* each leading to a DLT. Five additional patients were treated at dose level 2 without additional DLTs. At dose level 3, two patients had DLTs. One patient experienced grade 3 *lymphocyte count decreased* and grade 3 *neutrophil count decreased*. The other developed a grade 3 then eventually a grade 4 *neutrophil count decreased* in addition to a grade three *platelet count decreased,* thus, reaching the stopping number, and dose level 2 was determined to be the maximum tolerated dose (MTD). All DLT’s occurred in the final week of CRT. Of note, only one fraction of radiotherapy was not completed in the 11 patients. Temozolomide compliance was 95.2%, 90% and 90%, and Selinexor compliance was 100%, 91.7% and 83.3%, for dose levels 1, 2 and 3, respectively. One patient had a grade 3 RTOG CNS toxicity at 7 months, that resolved to a grade 2 without a change in therapy and was unrelated to progression.

**Table 2:**
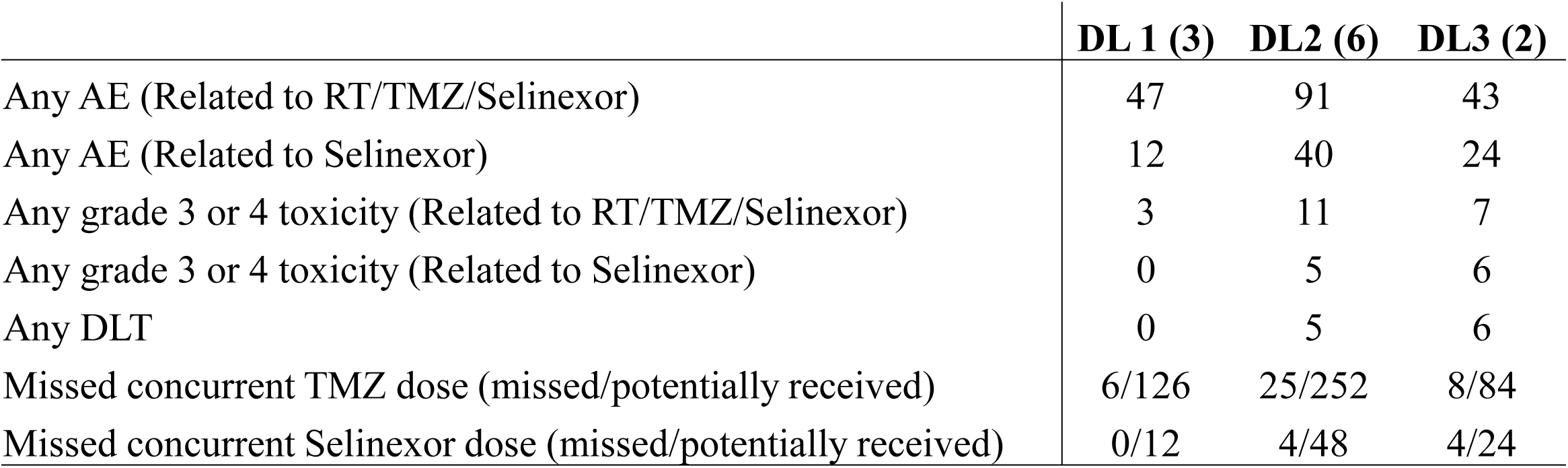
Protocol toxicities deemed definitely, possibly, or likely related to study treatment. DL = dose level, AE = adverse event, RT = radiotherapy, TMZ = temozolomide, DLT = dose limiting toxicity.

|  | DL 1 (3) | DL2 (6) | DL3 (2) |
| --- | --- | --- | --- |
| Any AE (Related to RT/TMZ/Selinexor) | 47 | 91 | 43 |
| Any AE (Related to Selinexor) | 12 | 40 | 24 |
| Any grade 3 or 4 toxicity (Related to RT/TMZ/Selinexor) | 3 | 11 | 7 |
| Any grade 3 or 4 toxicity (Related to Selinexor) | 0 | 5 | 6 |
| Any DLT | 0 | 5 | 6 |
| Missed concurrent TMZ dose (missed/potentially received) | 6/126 | 25/252 | 8/84 |
| Missed concurrent Selinexor dose (missed/potentially received) | 0/12 | 4/48 | 4/24 |

### Time to Progression and Response

Median OS was 17.4 months (95% CI: 14.1-NR(not reached)) and PFS was 15.9 months (95% CI: 6.2-28.5m)), as shown in **Figure 2**. However, several patients exhibited pseudoprogression (at 5, 9, 10 and 23 months), beyond the expected 12-week interval ^22^.Two patients were progression-free and alive at the time of the data freeze (March 2026). **Figure 3**, panel A, demonstrates an apparent tumour recurrence 5 months post-RT completion, however the patient was clinically asymptomatic. After consultation with the independent monitoring committee, we chose to follow the patient, who remains alive without evidence of progression at 65 months (final image of panel A). A second patient (panel B) had a similar radiographic finding and was taken to the OR for resection (at the patient’s behest) 23-month post-RT. Pathology from that specimen showed no evidence of tumour and the patient is 11 months post-op without evidence of disease (panel B final image). Six of the patients who failed had an MRI that was evaluable for the protocol derived pattern of failure (POF_p_). The location was defined as the percentage of the contrast enhanced tumour (CET) recurrence that occurred within the 95% (57 Gy) isodose line derived from the original treatment plan. Five patients had a *central* (POF_p_) (>95% of CET within the 57 Gy line) failure and one had a *distant* (POF_p_) (<20% of CET within the 57 Gy line). Following progression on our trial, patients received a mean of one additional treatment before death (range 0-2), with the 3 patients receiving bevacizumab or re-excision, and 1 receiving adjuvant reirradiation post-re-excision

**Figure 2:**
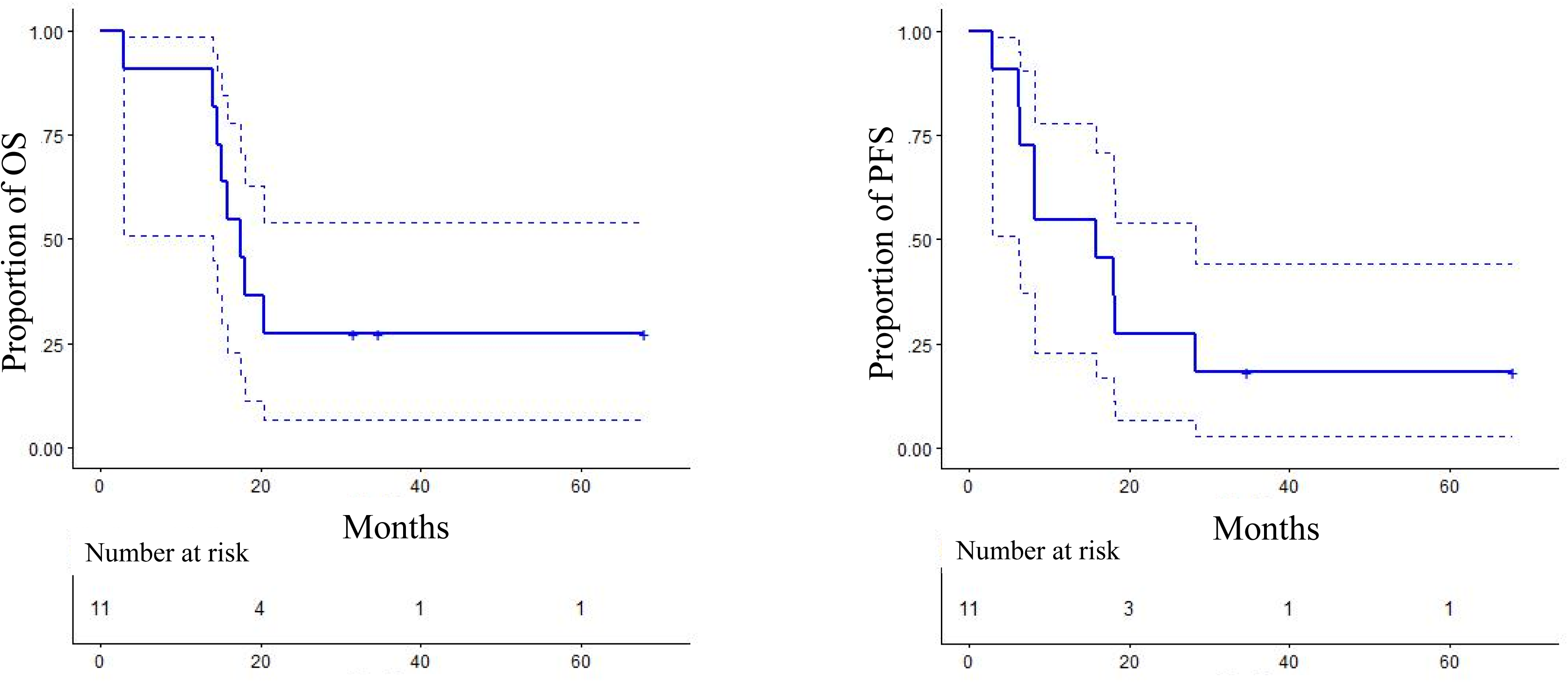
Kaplan-Meier analysis of overall survival (A) and progression-free survival (B). Dotted lines represent the 95% confidence limits. Blue cross hatches represent last follow-up dates for the patients who have not failed and are still alive.

**Figure 3:**
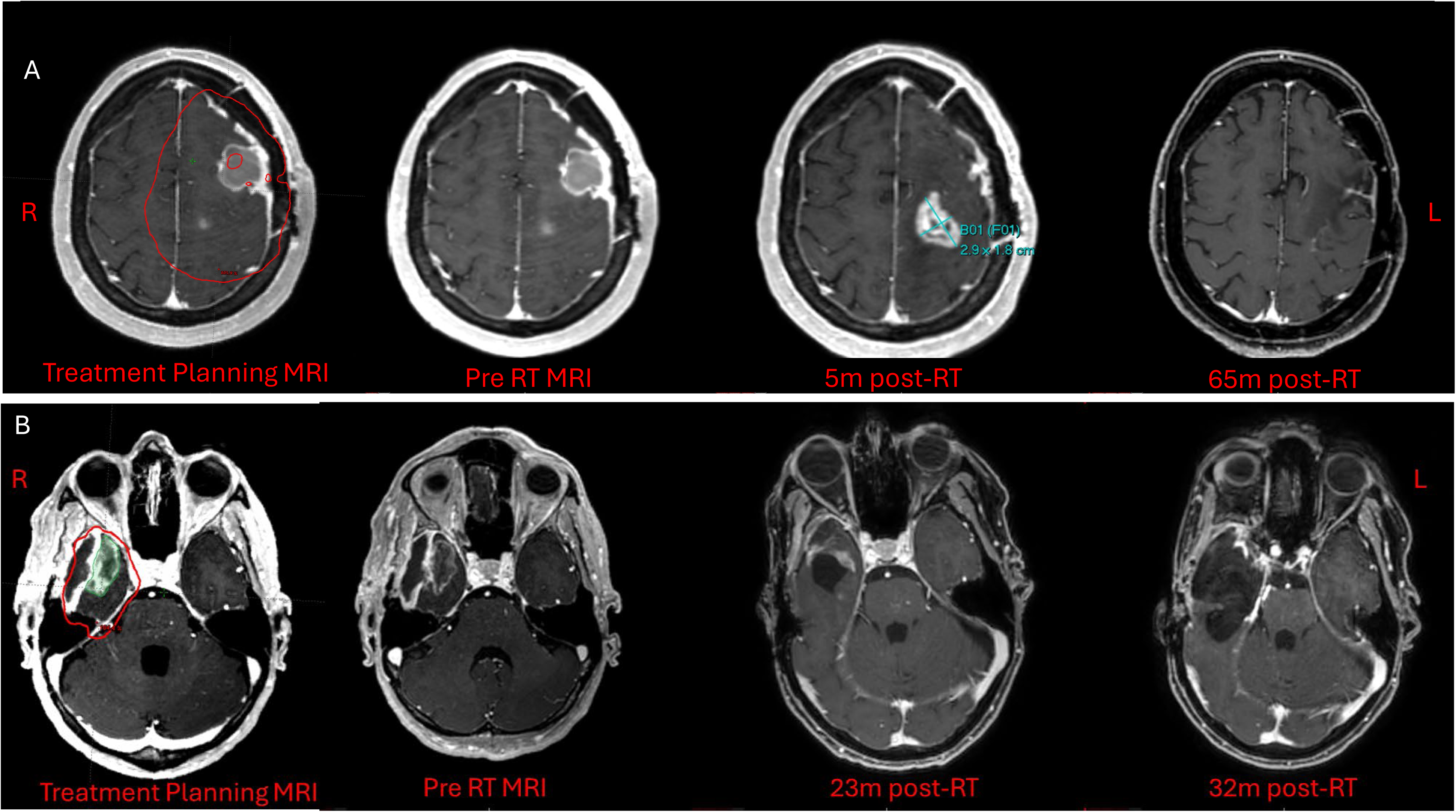
MR imaging of two examples of pseudoprogression at 5m (A) and at 23m (B) post-radiotherapy. The red line represents the 60 Gy dose line and the green area represents the GTV. R = right, L = left.

### Secondary Endpoints: Patient Reported Outcomes

At baseline, all patients were able to undergo symptom screening using the MDASI-BT questionnaire. Subsequently, 91%, 100%, 82%, and 82% filled out repeat questionnaires at the completion of treatment, 1 month, 3 months, and 5 months post-RT, respectively. The overall symptom scores were stable (**Figure 4),** with a small but not statistically significant increase in scores at the 1-month follow-up time point which receded at the 3-month follow-up. Three of the 11 patients had an increase in steroid use from the COT to the 5-month time point but did not correlate with an increase in their respective MDASI scores.

**Figure 4:**
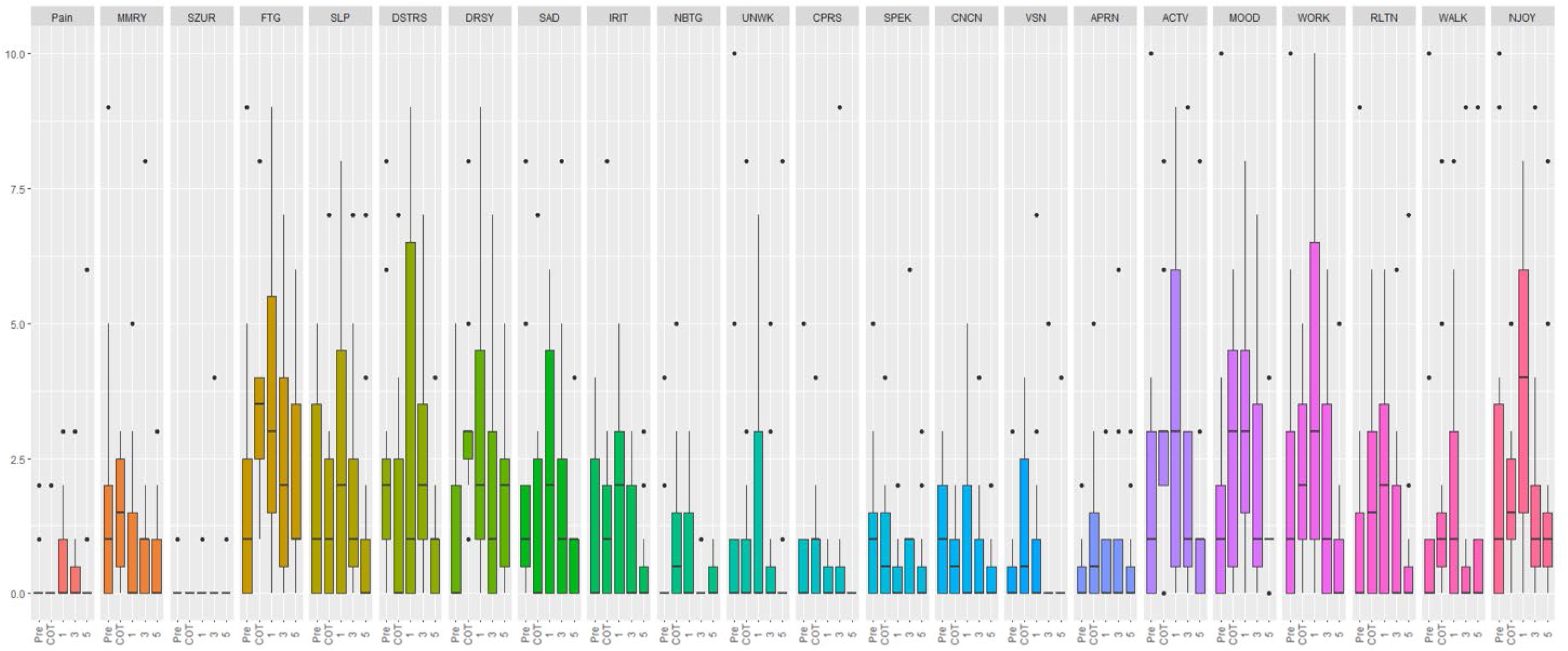
Box plots of MDASI-BT questionnaires across time. The upper and lower whiskers represent the range. The middle line box represents the 2nd quartile, while the lower and upper borders of the box represent the 1st and 3rd quartiles, respectively. Dots presented above or below box plots are points greater than 2 SD. Pain = Pain; MMRY =Memory Issues; SZUR =Seizures; FTG = Fatigue; SLP = Disturbed Sleep; DSTRS = Distress; DRSY = Drowsiness; SAD = Sadness; IRIT = Irritability; NBTG = Numbness/Tingling; UNWK = Unilateral Weakness; CPRS = Comprehension/Understanding; SPEK = Speaking Difficulty; CNCN = Concentration; VSN = Vision; APRN = Appearance; ACTV = General Activity; MOOD = Mood; WORK = Work Issues; RLTN = Relationship Issues; WALK = Walking; NJOY = Enjoyment.

## Discussion

In this phase I dose-escalation study evaluating the addition of Selinexor, used as a radiosensitizer, to standard radiotherapy and temozolomide we showed the combination was feasible and generally well tolerated in patients with newly diagnosed grade IV glioma. At the first two dose levels, treatment compliance was high, with minimal observable toxicities. At the final dose level both patients had DLTs that were attributable to Selinexor and the MTD was determined to be 60mg/m^2^ twice weekly, on weeks 1, 2, 4, 5 of chemoradiation. The late toxicity profile was similar to that commonly observed with standard chemoradiotherapy protocols (<10%). These findings support the safety of combining Selinexor with concurrent chemoradiation and establish a foundation for further dose escalation and efficacy evaluation.

Despite decades of investigation, efforts to improve outcomes in GBM through modification of radiation delivery or incorporation of radiosensitizers have largely been unsuccessful. While temozolomide remains the only agent to demonstrate a survival benefit in the upfront setting, its contribution as a radiosensitizer remains incompletely defined ^11^. Selinexor represents a mechanistically distinct approach, targeting molecules that are exported from the nucleus. Preclinical studies have suggested that inhibition of XPO1 enhances radiosensitivity, particularly in glioblastoma stem-like cells, which are thought to contribute to radioresistance and tumour recurrence ^21^. Our clinical findings are consistent with this data, demonstrating that this combination can be delivered safely in the upfront setting.

Although this study was not powered to assess efficacy, preliminary clinical outcomes appear encouraging. A subset of patients remains progression-free, and overall survival compares favorably to historical controls, though interpretation is limited by the small sample size and lack of a comparator arm ^10^. The median PFS of 15.9 months is twice the expected length (median 6-7 months) and may be due to the efficacy of Selinexor as a radiosensitizer. Further studies are needed to validate this effect. While early pseudoprogression generally suggests increased cell kill portending improved outcomes, the significance of delayed pseudoprogression remains unclear^23^. The apparent increase in late pseudoprogression in our cohort also raises the possibility that Selinexor may augment radiation-induced tumour and microenvironmental changes, potentially through enhanced DNA damage response inhibition or modulation of inflammatory signaling ^21^. This phenomenon, while potentially confounding radiographic interpretation, may also reflect enhanced local treatment effect. These findings emphasize the importance of cautious interpretation of early and intermediate post-treatment imaging and suggest that advanced imaging modalities or biomarker-driven approaches may be needed to better distinguish true progression from treatment effect in future studies incorporating Selinexor. The limitations of the present study are inherent to dose-escalation trials, which include a small sample size and lack of a control group. In addition, the protocol amendment introducing twice-weekly dosing introduces heterogeneity in treatment exposure across dose levels.

In conclusion, Selinexor can be safely combined with standard radiotherapy and temozolomide in patients with newly diagnosed GBM. The observed safety profile, along with signals of potential clinical activity and enhanced treatment effect, support further investigation in larger studies. Future trials should incorporate prospective evaluation of optimal dosing, integration with molecular stratification, and refined imaging criteria to better characterize response and distinguish pseudoprogression from true disease progression.

## Data Availability

Data Availability Statement for this Work: research data are stored in an institutional repository and will be shared upon request to the corresponding author.

---

**Supplemental Table 1:** Treatment planning volumes and volumes that received 60 Gy. GTV = gross tumour volume, PTV = planning target volume, CTV = clinical treatment volume, cm = centimeter.

| EORTC Volumes |  |
| --- | --- |
| GTV | Cavity + enhancing tumor |
| CTV | GTV + 2cm |
| PTV | CTV + 3-5mm |
| RTOG Boost Volumes |  |
| GTV | Cavity + enhancing tumor |
| CTV | GTV + 2cm |
| PTV | CTV + 3-5mm |
| Organ at Risk | Objective |
| Brainstem | Dmax < 60Gy |
| Chiasm | Dmax < 55Gy |
| Cochlea | mean < 45 Gy |
| Eyes | Macula < 45Gy |
| Optic nerves | Dmax < 55 Gy |
| RT Volumes to 60Gy |  |
| Median GTV, cc (range) | 24.2 (7.5-40.7) |
| Median CTV, cc (range) | 162.7 (75.7-300.2) |
| Median PTV, cc (range) | 222.3 (116.1-388.2) |

